## Supplemental Tables and FIgures for "Resurgence of malaria in Uganda despite sustained indoor residual spraying and repeated long lasting insecticidal net distributions"

**Supplemental Table 1.** Adjusted incident rate ratios and 95% for study objective 1.

| *Months since IRS was initiated* | *Incidence Rate Ratio* | *P-value* | *Lower bound of 95% CI* | *Upper bound of 95% CI* |
|---|---|---|---|---|
| *0* | REF | | | |
| *1* | 0.694 | 0.004 | 0.540 | 0.891 |
| *2* | 0.725 | 0.252 | 0.418 | 1.257 |
| *3* | 0.384 | 0.000 | 0.284 | 0.520 |
| *4* | 0.464 | 0.000 | 0.315 | 0.684 |
| *5* | 0.802 | 0.070 | 0.632 | 1.018 |
| *6* | 0.702 | 0.002 | 0.560 | 0.880 |
| *7* | 0.409 | 0.000 | 0.270 | 0.621 |
| *8* | 0.431 | 0.000 | 0.385 | 0.482 |
| *9* | 0.329 | 0.005 | 0.151 | 0.715 |
| *10* | 0.357 | 0.000 | 0.213 | 0.598 |
| *11* | 0.209 | 0.000 | 0.130 | 0.336 |
| *12* | 0.385 | 0.018 | 0.175 | 0.851 |
| *13* | 0.302 | 0.000 | 0.163 | 0.560 |
| *14* | 0.352 | 0.013 | 0.155 | 0.801 |
| *15* | 0.283 | 0.000 | 0.207 | 0.386 |
| *16* | 0.443 | 0.003 | 0.257 | 0.764 |
| *17* | 0.659 | 0.218 | 0.340 | 1.280 |
| *18* | 0.537 | 0.023 | 0.314 | 0.918 |
| *19* | 0.579 | 0.150 | 0.275 | 1.218 |
| *20* | 0.535 | 0.045 | 0.291 | 0.985 |
| *21* | 0.585 | 0.099 | 0.310 | 1.105 |
| *22* | 0.746 | 0.345 | 0.406 | 1.370 |
| *23* | 0.370 | 0.085 | 0.119 | 1.149 |
| *24* | 0.439 | 0.043 | 0.198 | 0.973 |
| *25* | 0.335 | 0.000 | 0.234 | 0.479 |
| *26* | 0.581 | 0.023 | 0.364 | 0.928 |
| *27* | 1.085 | 0.577 | 0.815 | 1.443 |
| *28* | 1.151 | 0.591 | 0.689 | 1.922 |
| *29* | 0.524 | 0.046 | 0.277 | 0.990 |
| *30* | 0.598 | 0.278 | 0.236 | 1.515 |
| *31* | 0.325 | 0.008 | 0.141 | 0.747 |
| *32* | 0.214 | 0.000 | 0.125 | 0.366 |
| *33* | 0.232 | 0.000 | 0.117 | 0.461 |
| *34* | 0.214 | 0.000 | 0.108 | 0.425 |
| *35* | 0.110 | 0.000 | 0.054 | 0.223 |
| *36* | 0.110 | 0.000 | 0.060 | 0.202 |
| *37* | 0.111 | 0.000 | 0.050 | 0.248 |
| *38* | 0.162 | 0.000 | 0.082 | 0.317 |
| *39* | 0.100 | 0.000 | 0.086 | 0.116 |
| *40* | 0.105 | 0.000 | 0.034 | 0.328 |
| *41* | 0.089 | 0.000 | 0.032 | 0.251 |
| *42* | 0.093 | 0.000 | 0.042 | 0.203 |
| *43* | 0.083 | 0.000 | 0.038 | 0.183 |
| *44* | 0.095 | 0.000 | 0.056 | 0.161 |

**Supplemental Table 1.** Adjusted incident rate ratios and 95% for study objective 1.

| *Months since IRS was initiated* | *Incidence Rate Ratio* | *P-value* | *Lower bound of 95% CI* | *Upper bound of 95% CI* |
|---|---|---|---|---|
| *45* | 0.128 | 0.000 | 0.060 | 0.274 |
| *46* | 0.075 | 0.000 | 0.036 | 0.157 |
| *47* | 0.067 | 0.000 | 0.043 | 0.103 |
| *48* | 0.100 | 0.000 | 0.057 | 0.174 |
| *49* | 0.109 | 0.000 | 0.065 | 0.181 |
| *50* | 0.134 | 0.000 | 0.052 | 0.348 |
| *51* | 0.140 | 0.000 | 0.085 | 0.230 |
| *52* | 0.118 | 0.000 | 0.090 | 0.154 |
| *53* | 0.222 | 0.000 | 0.147 | 0.336 |
| *54* | 0.261 | 0.000 | 0.170 | 0.399 |
| *55* | 0.276 | 0.000 | 0.160 | 0.478 |
| *56* | 0.291 | 0.000 | 0.227 | 0.374 |
| *57* | 0.232 | 0.000 | 0.161 | 0.334 |
| *58* | 0.281 | 0.000 | 0.147 | 0.536 |
| *59* | 0.355 | 0.028 | 0.141 | 0.892 |
| *60* | 0.554 | 0.245 | 0.205 | 1.499 |
| *61* | 0.867 | 0.761 | 0.344 | 2.185 |
| *62* | 0.924 | 0.762 | 0.552 | 1.546 |
| *63* | 0.459 | 0.095 | 0.183 | 1.146 |
| *64* | 0.400 | 0.010 | 0.199 | 0.803 |
| *65* | 0.288 | 0.000 | 0.171 | 0.483 |
| *66* | 0.349 | 0.000 | 0.267 | 0.455 |
| *67* | 0.403 | 0.000 | 0.276 | 0.587 |
| *68* | 0.570 | 0.000 | 0.422 | 0.769 |
| *69* | 0.474 | 0.000 | 0.313 | 0.718 |
| *70* | 0.670 | 0.015 | 0.486 | 0.925 |
| *71* | 1.084 | 0.572 | 0.819 | 1.435 |
| *72* | 0.729 | 0.257 | 0.423 | 1.258 |
| *73* | 1.138 | 0.585 | 0.715 | 1.812 |
| *74* | 1.639 | 0.119 | 0.881 | 3.049 |
| *75* | 1.595 | 0.236 | 0.736 | 3.456 |
| *76* | 1.238 | 0.507 | 0.659 | 2.324 |
| *77* | 0.683 | 0.075 | 0.449 | 1.039 |
| *78* | 1.278 | 0.512 | 0.614 | 2.659 |
| *79* | 1.529 | 0.223 | 0.773 | 3.026 |
| *80* | 1.610 | 0.140 | 0.856 | 3.028 |
| *81* | 1.253 | 0.608 | 0.529 | 2.964 |

Supplemental Figure 1. Adjusted IRR from multilevel negative binomial model comparing the period after IRS was initiated to the period before IRS was initiated with unadjusted case counts as model outcome. Vertical bars represent the 95% CI around adjusted IRR. Effect estimates in grey are published previously.

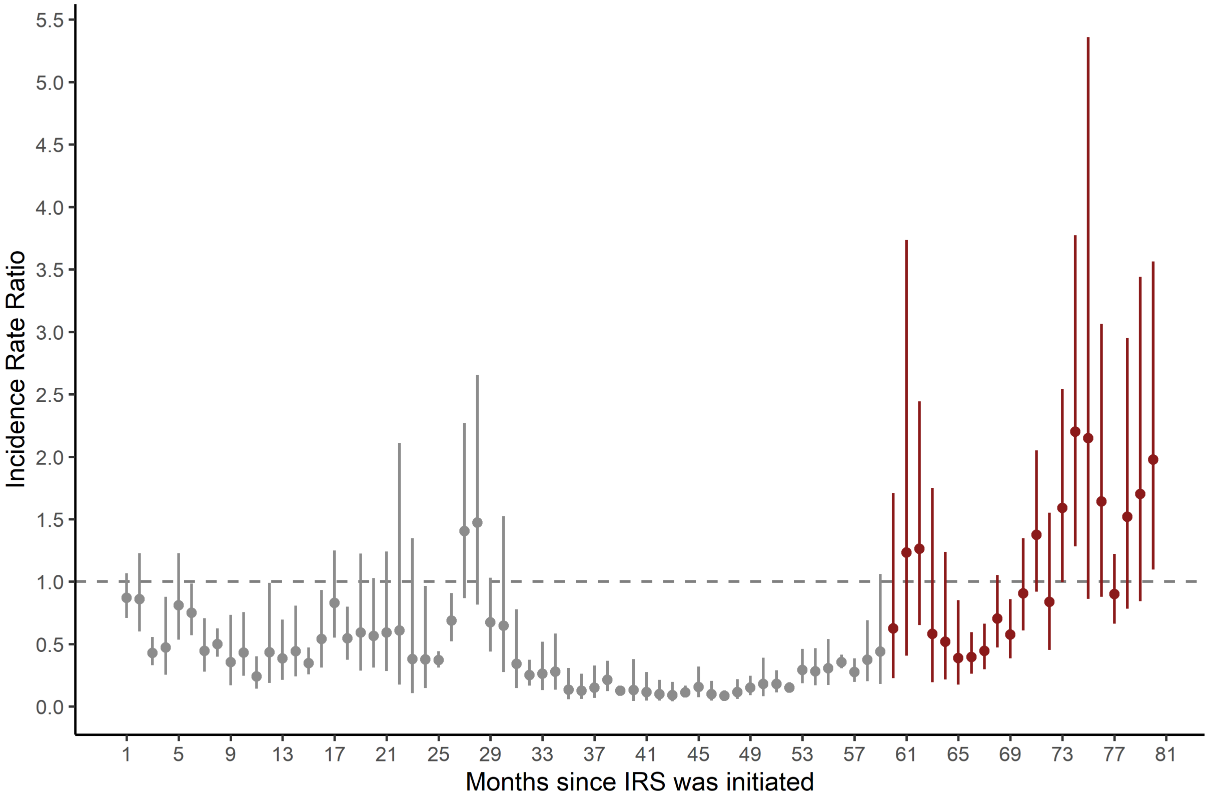

Supplemental Figure 2. Adjusted IRR from multilevel negative binomial model comparing the period after IRS was initiated to the period before IRS was initiated leaving out 2 sites that stopped IRS in 2021 (Orum and Alebtong). Vertical bars represent the 95% CI around adjusted IRR. Effect estimates in grey are published previously.

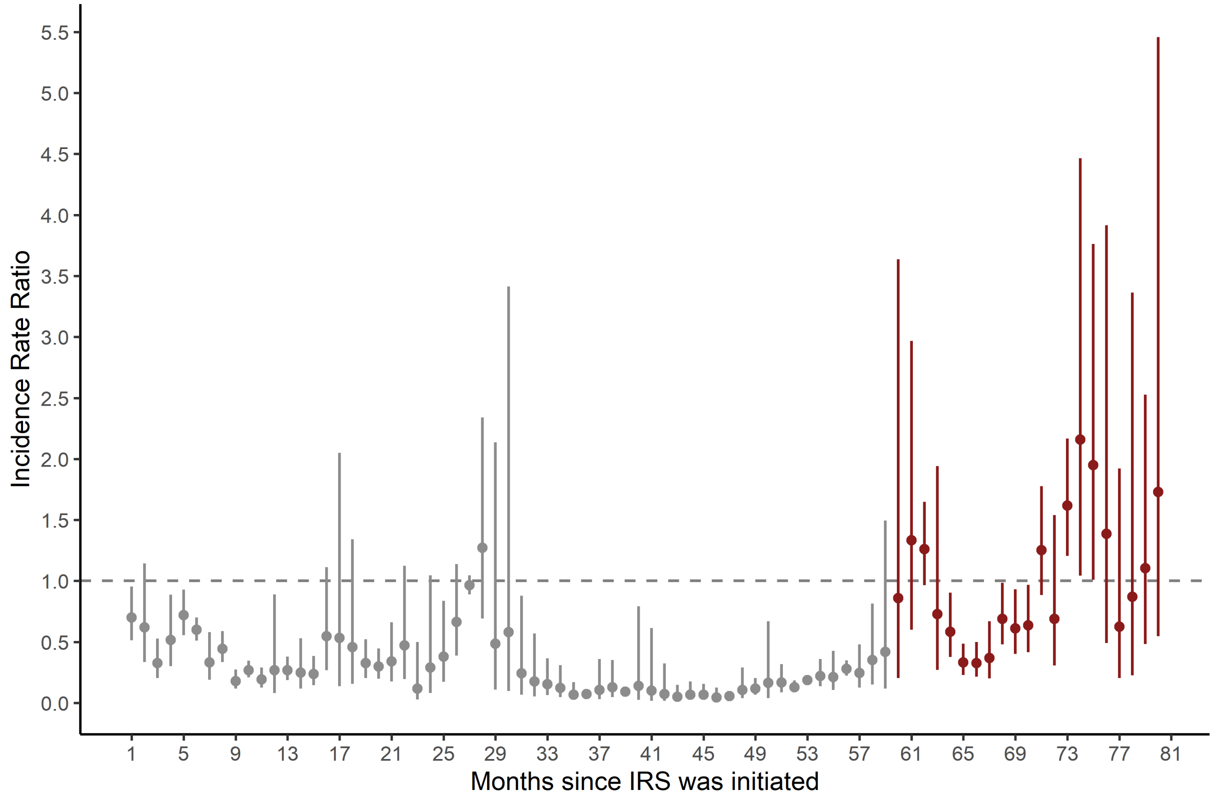

**Supplemental Table 2.** Adjusted incident rate ratios and 95% for study objective 2.

| *Month-Year* | Incident rate ratio IRS vs non-IRS | Lower bound 95% CI | Upper bound 95% CI |
|---|---|---|---|
| *Jan-20* | 0.35 | 0.19 | 0.66 |
| *Feb-20* | 0.39 | 0.21 | 0.71 |
| *Mar-20* | 0.29 | 0.16 | 0.53 |
| *Apr-20* | 0.22 | 0.12 | 0.40 |
| *May-20* | 0.26 | 0.14 | 0.48 |
| *Jun-20* | 0.21 | 0.12 | 0.38 |
| *Jul-20* | 0.18 | 0.10 | 0.33 |
| *Aug-20* | 0.23 | 0.12 | 0.42 |
| *Sep-20* | 0.27 | 0.15 | 0.50 |
| *Oct-20* | 0.32 | 0.17 | 0.60 |
| *Nov-20* | 0.47 | 0.25 | 0.87 |
| *Dec-20* | 0.83 | 0.45 | 1.52 |
| *Jan-21* | 0.70 | 0.38 | 1.30 |
| *Feb-21* | 0.93 | 0.50 | 1.72 |
| *Mar-21* | 1.22 | 0.66 | 2.25 |
| *Apr-21* | 1.31 | 0.71 | 2.41 |
| *May-21* | 1.38 | 0.76 | 2.51 |
| *Jun-21* | 1.25 | 0.68 | 2.30 |
| *Jul-21* | 1.42 | 0.77 | 2.61 |
| *Aug-21* | 1.66 | 0.91 | 3.04 |
| *Sep-21* | 1.95 | 1.07 | 3.54 |
| *Oct-21* | 1.60 | 0.89 | 2.88 |

Supplemental Figure 3. Adjusted IRR comparing IRS sites to non-IRS sites over the study period with unadjusted case counts as model outcome. Vertical bars represent the 95% CI around adjusted IRR.

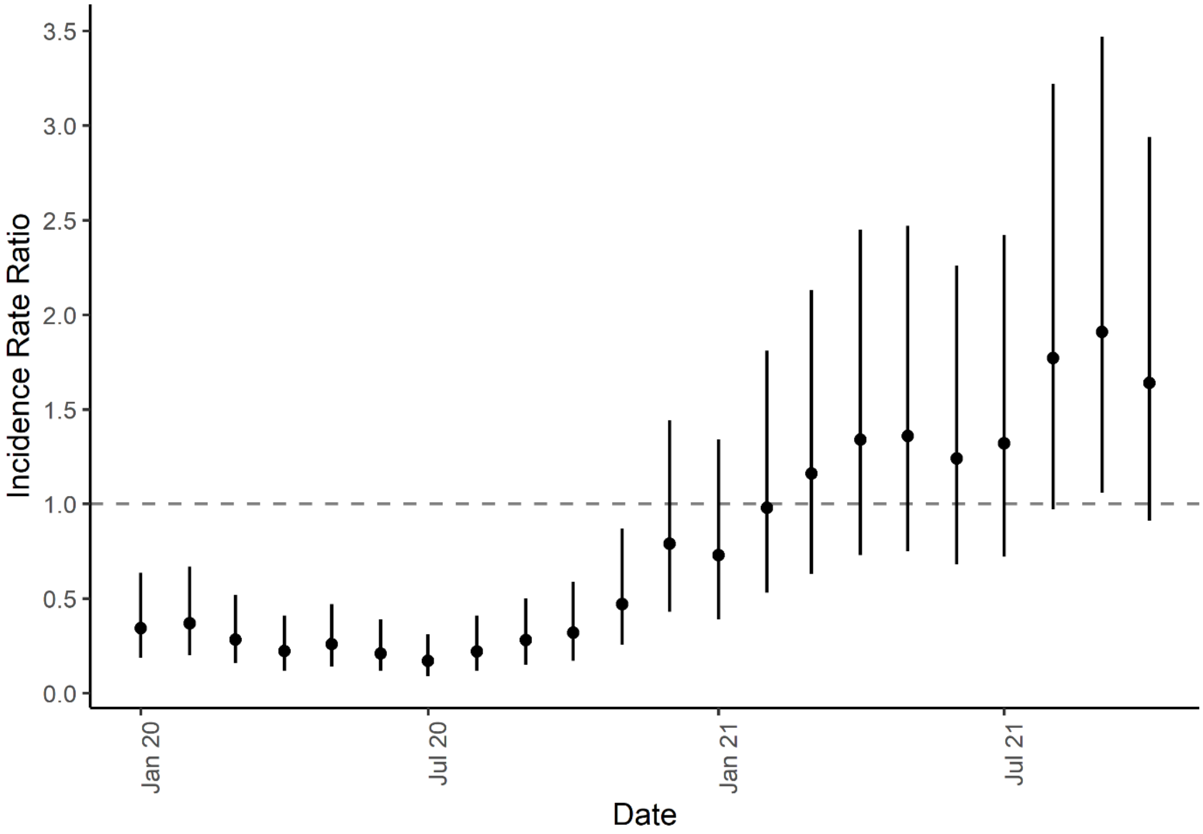

**Supplemental Table 3. Timing and formulation IRS campaigns.**

| MRC | District | Details of IRS campaign | 1st round (Coverage) | 2nd round (Coverage) | 3rd round (Coverage) | 4th round (Coverage) | 5th round (Coverage) | 6th round (Coverage) | 7th round (Coverage) | 8th round (Coverage) | 9th round (Coverage) |
|---|---|---|---|---|---|---|---|---|---|---|---|
| Nagongera HCIV | Tororo | Date | 08-Dec-14 to 19-Feb-15 | 08-Jun-15 to 12-Jul-15 | 02-Nov-15 to 12-Dec-15 | 12-Jun-16 to 9-Jul-16 | 17-July-17 to 19-Aug-17 | 11-Jun-18 to 27-7-18 | 18-Mar-19 to 15-Apr-19 | 2-Mar-20 to 28-Mar-2020 | 1-Mar-20 to 26-Mar-2021 |
| | | Formulation | Bendiocarb | Bendiocarb | Bendiocarb | Actelic | Actellic | Actellic | Actellic | Fludora Fusion WP-SB* | Fludora Fusion WP-SB |
| | | Coverage | Not available | (94.60%) | (96.30%) | (93.0%) | (94.90%) | (95.80%) | (92.10%) | (93.20%) | (99.4%) |
| Amolatar HCIV | Amolatar | Date | 08-Dec-14 to 19-Feb-15 | 08-Jun-15 to 12-Jul-15 | 12-Oct-15 to 07-Nov-15 | 24-Oct-16 to 19-Nov-16 | 2-May-17 to 6-June-17 | 9-Apr-18 to 12-May-18 | 27-May-19 to 27-June-19 | 25-May-2020 to 20-Jun-2020 | 26 April to 20 May 2021 |
| | | Formulation | Bendiocarb | Bendiocarb | Bendiocarb | Bendiocarb | Actellic | Actellic | Actellic | Fludora Fusion WP-SB† | Fludora Fusion WP-SB |
| | | Coverage | Not available | (96.10%) | (91.40%) | (94.1%) | (95.80%) | (96.10%) | (94.20%) | (96.20%) | (91.5% ) |
| Dokolo HCIV | Dokolo | Date | 08-Dec-14 to 19-Feb-15 | 08-Jun-15 to 12-Jul-15 | 12-Oct-15 to 07-Nov-15 | 18-Apr-16 to 9-Jul-16 | 2-May-17 to 6-June-17 | 9-Apr-18 to 12-May-18 | 27-May-19 to 27-June-19 | 25-May-2020 to 20-Jun-2020 | 26 April to 25 May 2021 |
| | | Formulation | Bendiocarb | Bendiocarb | Bendiocarb | Actellic | Actellic | Actellic | Sumishield 50W | Fludora Fusion WP-SB† | Fludora Fusion WP-SB |
| | | Coverage | Not available | (88.70%) | (92.10%) | (94.80%) | (97.60%) | (94.30%) | (95.20%) | (94.60%) | (91.0% ) |
| Orum HCIV | Otuke | Date | 08-Dec-14 to 19-Feb-15 | 08-Jun-15 to 12-Jul-15 | 02-Nov-15 to 12-Dec-15 | 18-Apr-16 to 9-Jul-16 | 17-July-17 to 19-Aug-17 | 11-Jun-18 to 27-7-18 | 27-May-19 to 27-June-19 | 25-May-2020 to 20-Jun-2020 | |
| | | Formulation | Bendiocarb | Bendiocarb | Bendiocarb | Actellic | Actellic | Actellic | Actellic | Fludora Fusion WP-SB† | |
| | | Coverage | Not available | (96.60%) | (94.80%) | (97.60%) | (96.70%) | (98.40%) | (98.80%) | (96.50%) | |
| Alebtong HCIV | Alebtong | Date | 08-Dec-14 to 19-Feb-15 | 08-Jun-15 to 12-Jul-15 | 12-Oct-15 to 07-Nov-15 | 24-Oct-16 to 19-Nov-16 | 2-May-17 to 6-June-17 | 9-Apr-18 to 12-May-18 | 27-May-19 to 27-June-19 | 25-May-2020 to 20-Jun-2020 | |
| | | Formulation | Bendiocarb | Bendiocarb | Bendiocarb | Bendiocarb | Actellic | Actellic | Actellic | Fludora Fusion WP-SB† | |
| | | Coverage | Not available | (96.90%) | (95.40%) | (97.50%) | (98.30%) | (90.90%) | (95.00%) | (95.50%) | |

*2% of households received Actellic

†2% of households received Sumishield 50W
